# PATIENT PROFILING AND OUTCOME OF ANTIHYPERTENSIVE TREATMENT. An Exploratory Analysis of the VALUE Trial

**DOI:** 10.64898/2026.09.18.26363450

**Authors:** Peter W de Leeuw, Camilla L Søraas, Giuseppe Mancia, Maria H Mehlum, Kenneth Jamerson, Solko W Schalm, Sverre E Kjeldsen, Michael A Weber

## Abstract

**Background:** Randomized outcome trials have shown the benefit of antihypertensive treatment, but subgroup analyses exploring treatment responses in relation to patient characteristics are usually based on single factors rather than comprehensive patient profiles. We hypothesized that a combination of individual predictors better identifies patients who are more or less likely to achieve blood pressure (BP) control.

**Methods:** To test our hypothesis, we reanalysed the data from the double-blinded VALUE-outcome trial (n=15,313). The patient population was divided into an exploratory cohort (2/3) and a validation cohort (1/3). We constructed composite patient profiles by combining four proven predictors of BP control: age, severity of hypertension, comorbidity and previous treatment status. Logistic regression and Cox proportional hazard models were used to test whether BP control and cardiovascular event rates differed among these profiles, and whether profiling conferred additional predictive information beyond the individual factors alone.

**Results:** BP control rates differed significantly between patient profiles, regardless of treatment (likelihood ratio test, p<0.001). Results from the exploratory cohort were reproducible in the validation cohort. Profiling predicted BP control significantly better than any individual factor (p<0.001 for all comparisons). In addition, there were significant associations between profiles and the incidence of cardiovascular events. Adverse events, however, were not related to profiles.

**Conclusions:** We conclude that composite patient profiles may predict BP responses to antihypertensive treatment more accurately than single factors. These findings may help to better individualize antihypertensive therapy.

## Introduction

BP (BP) responses to widely used antihypertensive drugs show substantial heterogeneity among individual patients.^1^ In most studies, potential predictors of treatment responses have only been tested as separate factors, e.g. male versus female sex or presence versus absence of diabetes. If we want to implement a more personalized form of antihypertensive treatment, though, we need to better understand how more comprehensive patient profiles affect individual responses to antihypertensive agents. Knowing beforehand what the chances of treatment success are with different drugs in patients with different profiles could be particularly useful in general practice and potentially lead to greater treatment success with less medication and less consultations.

Properly executed post-hoc analyses of existing trials may provide the data which are necessary to explore the relationship between various composite patient profiles and the effect of treatment.^2^ Indeed, the recent analysis of the INSIGHT study showed that the odds of reaching a systolic BP below 140 mmHg during the first six months of treatment correlated significantly with composite patient profiles that were based on prognostic factors such as age, severity of hypertension, comorbidity and previous treatment status.^3^ However, that analysis did not report outcome data, so it remains uncertain whether such composite profiles are also associated with the occurrence of cardiovascular events. This is unfortunate as reduction of the latter is the ultimate goal of antihypertensive treatment. The VALUE trial, on the other hand, emphasized the prognostic importance of prompt BP control in hypertensive individuals at high cardiovascular risk but did not identify which specific patient profiles were associated with a better outcome.^4,5^ Given the size of the VALUE trial, a closer look at the data should enable us to validate the predictive value of composite patient profiles for early BP control with other antihypertensives and to assess the potential of such profiles to predict major outcomes. With this in mind, we undertook a post-hoc analysis of the VALUE trial with the following key question: can the findings of the INSIGHT study with respect to the prognostic value of patient profiles be confirmed and expanded with long-term outcomes of cardiovascular complications?

## Methods

### Trial description

The VALUE trial was a randomized, double-blind, parallel-group study in 15.245 hypertensive patients, aged 50 years or older with high cardiovascular risk^4^. Patients were allocated to treatment with either amlodipine or valsartan and the primary endpoint was a composite of cardiac mortality and morbidity. The results showed that reaching BP control, defined as a systolic BP below 140 mmHg, by six months was associated with significant benefits for subsequent major outcomes, irrespective of drug type. For the present analysis of the VALUE trial, we used the BP data at baseline and after six months of treatment to assess *early* BP control. The last recorded BP, i.e. just prior to an event or at censoring, were taken to determine *final* BP control. In addition, we assessed the incidence of the primary endpoint in relation to patient profiles. Patients without an event were censored at their last follow-up visit.

### Patient profiles

For the construction of composite patient profiles, we divided the participants of the trial in categories based on the variables age (<60, 60-80 and ≥ 80 years), baseline level of systolic BP (<160 or ≥160 mmHg), presence of organ damage (yes/no), comorbidity (none, obesity, diabetes alone or diabetes with obesity), and previous treatment status (no prior therapy, prior monotherapy or prior treatment with two or more drugs). These variables were chosen a priori because they have proven to be of prognostic significance in the treatment of hypertension.^6^ The presence of organ damage (cerebral, cardiac, renal and peripheral vascular) was defined as described in the original protocol of the study.^7^ Combining the four categories yields 3 × 2 × 2 × 4 × 3=144 possible patient profiles (see Supplemental file 1).

However, to ensure a reasonable estimate of treatment effects, we restricted our analysis to those profiles for which at least 50 participants were available. Individual patients for whom data essential for patient profiling were lacking were excluded from the analysis.

### Statistical analysis

We started our analysis by dividing the study population, in a 2:1 ratio and balanced for treatment and distribution of profiles, into two random samples. The larger fraction served to identify associations (exploratory cohort), and the other one to assess the validity of the results (validation cohort). Outcomes were BP control at six months, BP control just before an event or at the end of follow-up, first cardiovascular event and adverse events, all defined as binary variables (yes/no).

Using logistic regression analysis with the Wald chi-square statistic, we first assessed in the exploratory cohort, and for both treatments separately, the association between each patient profile and BP outcomes or events. Patient profiles were treated as categorical variables and compared to the weighted grand mean outcome in the specified group as reference (weighted effect coding models). Whenever appropriate, we applied Oldham’s correction to the baseline BP to avoid spurious relationships with changes in BP.^8,9^ To evaluate whether the association of patient profiles with BP effects differed between the two treatments, a combined logistic regression model was fitted which included both treatment groups and an interaction term. In case of no statistically significant difference, we also analysed the data of both groups together, using treatment as covariate rather than as a dividing variable. This same was done with sex as covariate.

We used linear regression and Bland-Altman analyses to test whether the results obtained in the exploratory cohort could be corroborated in the validation cohort.

Next, we assessed for both treatments the association between profile categories and clinical outcome (time to first event) using a Cox proportional hazard model with sex as covariate. Finally, we evaluated the association between profiles and adverse events.

Continuous data are expressed as medians with standard deviations and categorical data as percentages with 95% confidence intervals (95% CI). Associations are presented as odds ratios (ORs) with 95% CI as derived from the logistic regression models or, in case of events, as hazard ratios (HRs) with 95% CI. To avoid chance findings as may occur with multiple testing, all p-values were adjusted using Hommel’s method.^10^ Statistical significance was set at an adjusted p-value < 0.05. We used Stata software (version 19.5 with Python integration) and GraphPad Prism (version 11.0) for all statistical and graphical analyses.

## Results

Of the 144 profiles that are theoretically possible, 121 could be identified in the entire VALUE population. However, most of these contained only a few patients. When restricted to the exploratory cohort, regardless of treatment, 36 profiles consisted of 50 or more patients.

Further subdivision by treatment arm, resulted in 24 patient profiles which qualified for the present analysis (Table 1). Although the number of patients in each profile varied substantially, they were comparable among the two treatment groups.

**Table 1.** Distribution of patient profiles with 50 or more observations in both treatment groups.

| Profile number | Age | SBP | OD | Comorbidity | Previous treatment | Amlodipine (n) | Valsartan (n) |
| --- | --- | --- | --- | --- | --- | --- | --- |
| 14 | < 60 | <160 | Yes | None | Monotherapy | 193 | 201 |
| 17 | < 60 | <160 | Yes | Obesity | Monotherapy | 108 | 108 |
| 20 | < 60 | <160 | Yes | Diabetes | Monotherapy | 54 | 66 |
| 23 | < 60 | <160 | Yes | Obesity and diabetes | Monotherapy | 77 | 74 |
| 38 | < 60 | ≥160 | Yes | None | Monotherapy | 76 | 73 |
| 41 | < 60 | ≥160 | Yes | Obesity | Monotherapy | 50 | 50 |
| 50 | 60-80 | <160 | No | None | Monotherapy | 58 | 67 |
| 56 | 60-80 | <160 | No | Diabetes | Monotherapy | 60 | 50 |
| 62 | 60-80 | <160 | Yes | None | Monotherapy | 761 | 765 |
| 63 | 60-80 | <160 | Yes | None | Dual/triple therapy | 147 | 165 |
| 65 | 60-80 | <160 | Yes | Obesity | Monotherapy | 267 | 276 |
| 66 | 60-80 | <160 | Yes | Obesity | Dual/triple therapy | 69 | 74 |
| 68 | 60-80 | <160 | Yes | Diabetes | Monotherapy | 263 | 247 |
| 69 | 60-80 | <160 | Yes | Diabetes | Dual/triple therapy | 68 | 65 |
| 71 | 60-80 | <160 | Yes | Obesity and diabetes | Monotherapy | 209 | 194 |
| 72 | 60-80 | <160 | Yes | Obesity and diabetes | Dual/triple therapy | 60 | 70 |
| 85 | 60-80 | ≥160 | Yes | None | Naïve | 100 | 99 |
| 86 | 60-80 | ≥160 | Yes | None | Monotherapy | 448 | 411 |
| 87 | 60-80 | ≥160 | Yes | None | Dual/triple therapy | 92 | 100 |
| 89 | 60-80 | ≥160 | Yes | Obesity | Monotherapy | 128 | 143 |
| 92 | 60-80 | ≥160 | Yes | Diabetes | Monotherapy | 205 | 221 |
| 95 | 60-80 | ≥160 | Yes | Obesity and diabetes | Monotherapy | 155 | 117 |
| 110 | ≥80 | <160 | Yes | None | Monotherapy | 52 | 62 |
| 134 | ≥80 | ≥160 | Yes | None | Monotherapy | 50 | 51 |
| **Total** |  |  |  |  |  | **3750** | **3749** |

SBP: systolic blood pressure; OD: organ damage.

### Profile-related BP outcomes in the exploratory cohort

Baseline characteristics of the patients are presented in Table 2. At six months, a systolic BP below 140 mmHg was reached in 52% (95% CI: 50-53%) of the patients in the amlodipine arm and in 47% (95% CI: 46-48%) of those in the valsartan arm. This difference is statistically significant (p<0.001). However, as depicted in Figures 1 and 2, the probability of reaching BP control and the observed proportions of BP control varied from about 20 to 75% across the different profiles. Logistic regression analysis shows that overall, both in the amlodipine and in the valsartan group, patient profiles are significantly associated with BP control (likelihood ratio test, p<0.001 for both groups).

**Figure 1.**
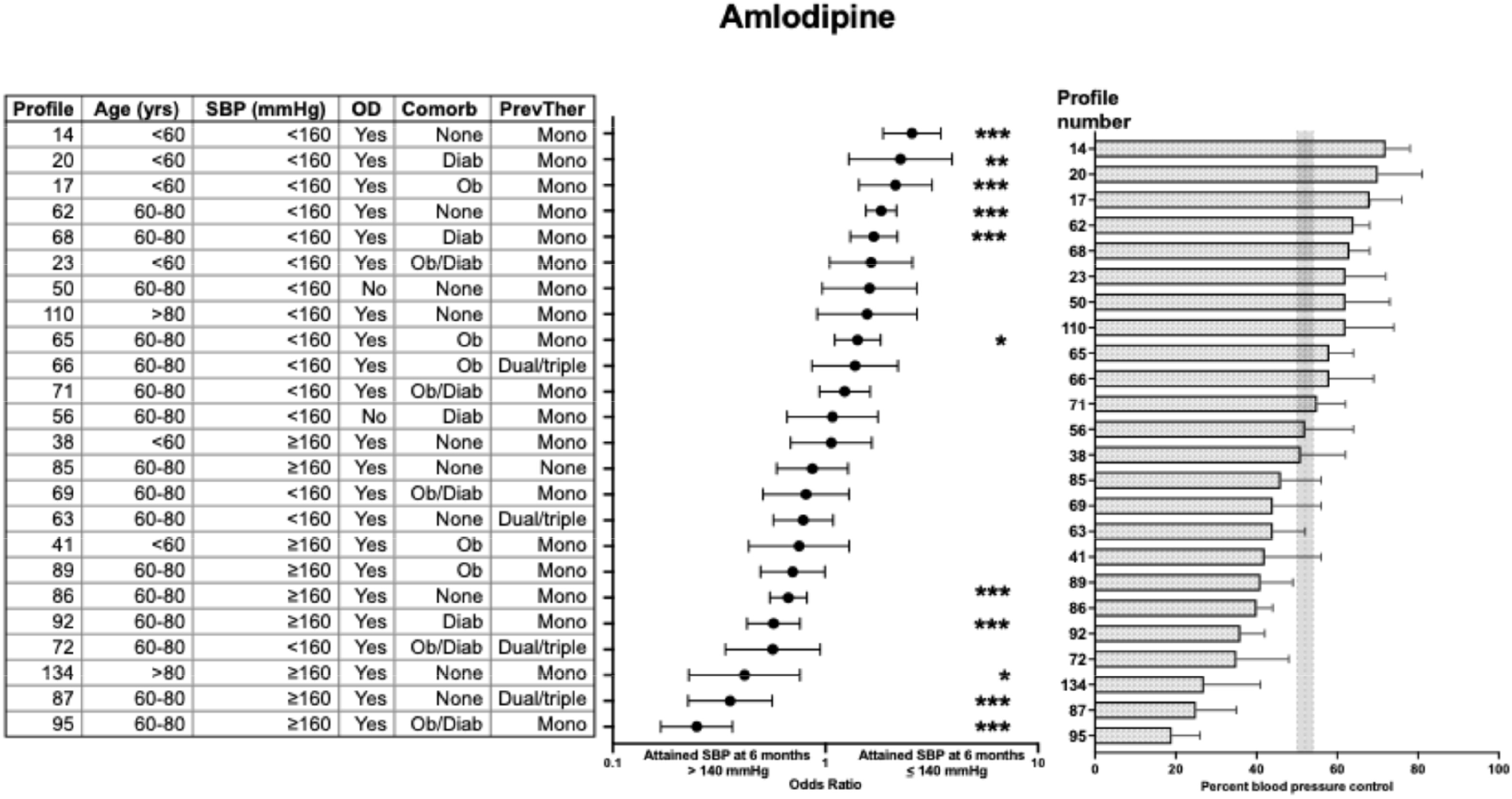
Blood pressure control in the amlodipine arm of the exploratory cohort. Left panel: the constituents of the patient profiles: profile number, age, systolic blood pressure (SBP), presence or absence of organ damage (OD), comorbidity (Comorb) and previous therapy (PrevTher). Diab: diabetes; Ob:obesity; Mono: monotherapy; Dual/triple: dual or triple therapy. Middle panel: odds ratios of achieving blood pressure control in each profile relative to that in the entire treatment group. Right panel: corresponding percentages of patients with blood pressure control for each profile (shaded area indicates average with 95% confidence interval for entire group). Data have been arranged in ascending order of odds ratio. Profiles 23, 72 and 89 were statistically significant in crude analysis but not anymore after adjustment for multiple testing. * p=0.01; ** p<0.01; *** p<0.001 (all p-values have been adjusted for multiple testing).

**Figure 2.**
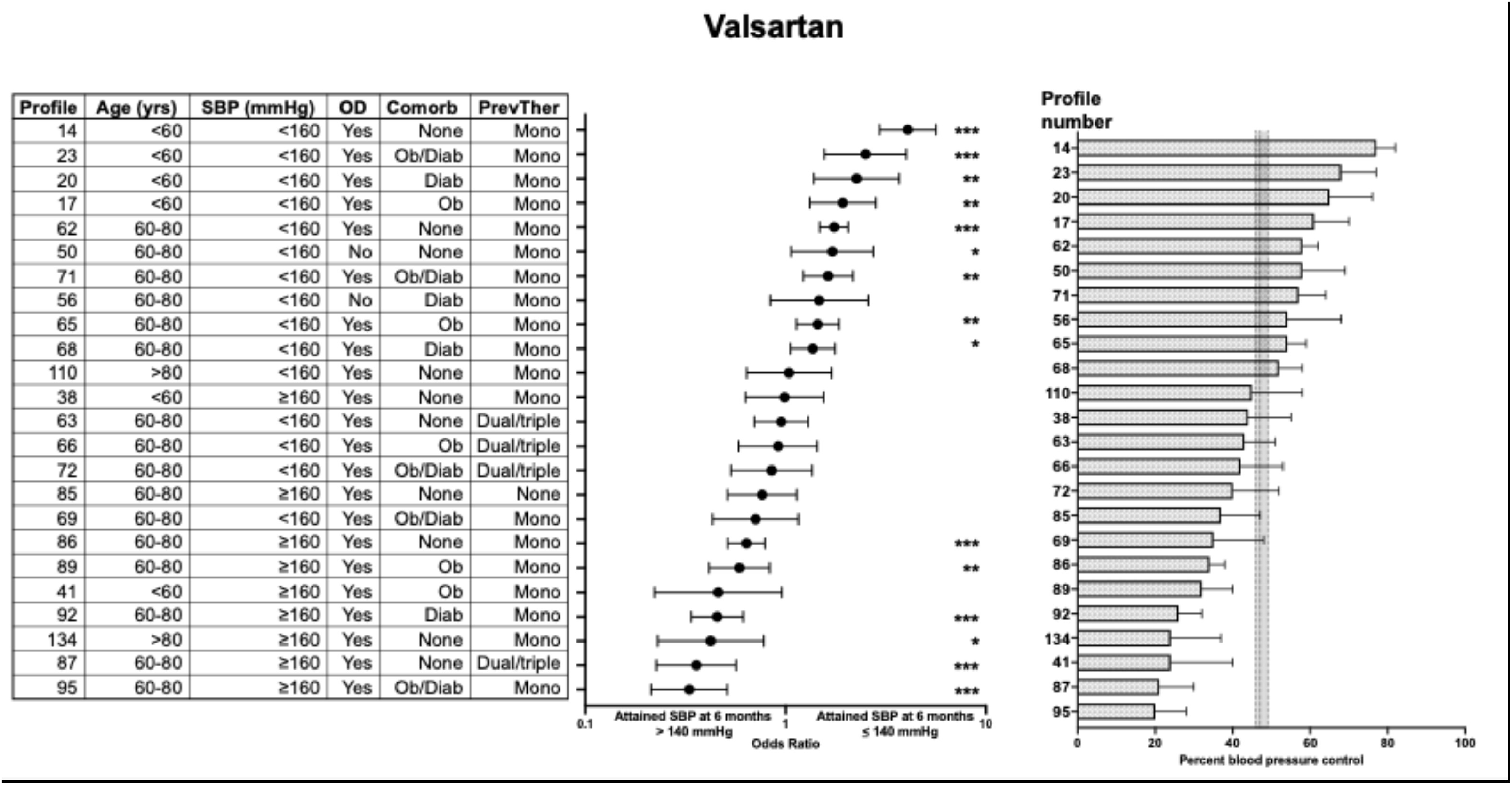
Blood pressure control in the valsartan arm of the exploratory cohort. Left panel: the constituents of the patient profiles: profile number, age, systolic blood pressure (SBP), presence or absence of organ damage (OD), comorbidity (Comorb) and previous therapy (PrevTher). Diab: diabetes; Ob:obesity; Mono: monotherapy; Dual/triple: dual or triple therapy. Middle panel: odds ratios of achieving blood pressure control in each profile relative to that in the entire treatment group. Right panel: corresponding percentages of patients with blood pressure control for each profile (shaded area indicates average with 95% confidence interval for entire group). Data have been arranged in ascending order of odds ratio. Profile 41 was statistically significant in crude analysis but not anymore after adjustment for multiple testing. * p<0.05; ** p<0.01; *** p<0.001 (all p-values have been adjusted for multiple testing).

**Table 2.** Baseline characteristics of the patients included in the analysis.

|  | Amlodipine |  | Valsartan |  |
| --- | --- | --- | --- | --- |
|  | Exploratory cohort | Validation cohort | Exploratory cohort | Validation cohort |
| N | 3750 | 1850 | 3749 | 1828 |
| Age (years) | 67 ± 8 | 67 ± 8 | 67 ± 8 | 67 ± 8 |
| Sex (Male/Female, %) | 58/42 | 57/43 | 57/43 | 60/40 |
| Baseline SBP (mmHg) | 153 ± 18 | 153 ± 19 | 152 ± 18 | 152 ± 19 |
| Target organ damage (n, %) | 3638 (97%) | 1793 (97%) | 3637 (97%) | 1773 (97%) |
| Obesity (n, %) | 1125 (30%) | 553 (30%) | 1095 (29%) | 534 (29%) |
| Diabetes (n, %) | 1151 (31%) | 568 (31%) | 1106(30%) | 536 (29%) |
| Treatment-naïve (n, %) | 101 (3%) | 48 (3%) | 101 (3%) | 49 (3%) |

SBP: systolic blood pressure.

Comparable significant associations exist when the achieved BP as a continuous variable is considered, both in those who did not reach the target BP and in those who did (p<0.001 for all). In other words, patients with less favourable profiles had a higher systolic BP at 6 months, even when this was below 140 mmHg, than patients with more advantageous profiles. In line with this, the latter needed less additional medications (e.g. beta-blockers, diuretics) than the former.

When the association of patient profiles with BP control at the last follow-up before an event or at censoring was examined, the same associations with profile categories were found (likelihood ratio test, p<0.001). At that time, 66% (95% CI: 65-68%) of the patients on amlodipine and 61% (95%CI: 59-62%) of those on valsartan had attained BP control (p<0.001).

Crude analyses showed that in 14 of the 24 profiles from the amlodipine group and 16 of the 24 profiles from the valsartan group, the percentage BP control differed significantly from the mean of their respective treatment groups. After Hommel’s correction for multiple testing, 11 and 15 associations respectively remained significant. Adjustment for sex as a covariate did not alter any of these results. Five profiles (14, 17, 20, 62 and 68) were associated with a higher-than-average likelihood of achieving BP control within six months in both treatment arms. Conversely, four profiles (87, 92, 95 and 134) consistently exhibited a lower likelihood of BP control.

Interaction analysis indicated that the relationship between profiles and BP control did not differ between the two treatment groups (p=0.67). Therefore, we repeated the analyses in the whole group of patients with treatment as covariate. This resulted in a greater number of significant associations (Table 3). Altogether, 11 profiles showed less-than-average BP responses.

**Table 3.** Odds ratios (with 95% confidence intervals) for attaining blood pressure control in relation to profile categories. Exploratory cohort only with both treatment groups combined; all data adjusted for sex.

| Profile | OR | p (crude) | p (Hommel) |
| --- | --- | --- | --- |
| 14 | 2.96 (2.37-3.69) | **<0.001** | **<0.001** |
| 17 | 1.85 (1.40-2.44) | **<0.001** | **<0.001** |
| 20 | 2.11 (1.44-3.08) | **<0.001** | **<0.001** |
| 23 | 1.86 (1.33-2.58) | **<0.001** | **<0.001** |
| 38 | 0.92 (0.67-1.26) | 0.59 | 0.62 |
| 41 | 0.53 (0.34-0.82) | **0.005** | **0.008** |
| 50 | 1.56 (1.09-2.23) | **0.01** | **0.02** |
| 56 | 1.13 (0.77-1.65) | 0.53 | 0.57 |
| 62 | 1.62 (1.47-1.77) | **<0.001** | **<0.001** |
| 63 | 0.78 (0.63-0.97) | **0.03** | **0.04** |
| 65 | 1.30 (1.11-1.54) | **0.002** | **0.003** |
| 66 | 1.01 (0.73-1.40) | 0.95 | 0.95 |
| 68 | 1.38 (1.16-1.64) | **<0.001** | **<0.001** |
| 69 | 0.67 (0.48-0.95) | **0.02** | **0.03** |
| 71 | 1.29 (1.06-1.56) | **0.001** | **0.01** |
| 72 | 0.63 (0.44-0.89) | **0.01** | **0.01** |
| 85 | 0.73 (0.56-0.97) | **0.03** | **0.04** |
| 86 | 0.59 (0.52-0.67) | **<0.001** | **<0.001** |
| 87 | 0.31 (0.22-0.43) | **<0.001** | **<0.001** |
| 89 | 0.58 (0.45-0.74) | **<0.001** | **<0.001** |
| 92 | 0.46 (0.37-0.56) | **<0.001** | **<0.001** |
| 95 | 0.24 (0.18-0.32) | **<0.001** | **<0.001** |
| 110 | 1.15 (0.80-1.66) | 0.45 | 0.52 |
| 134 | 0.35 (0.23-0.56) | **<0.001** | **<0.001** |

OR: odds ratio. Significant effects indicated in bold. P (crude): p-value in crude analysis. P (Hommel): p-value after Hommel's correction for multiple testing.

### Validation cohort

The proportion of patients from the validation cohort who achieved BP control at 6 months was 51% (95% CI:49-53%) in the amlodipine group and 48% (95% CI: 47-50%) in the valsartan group. When stratified by treatment group, consistent associations were observed among the exploratory and the validation cohort. Again, there were differences between the various profile categories in the validation cohort, but patterns aligned well with those in the exploratory cohort. This was true not only for BP at 6 months, but also for end-of-treatment BP. Figure 3 displays the relationships between the predicted BP control rates as derived from the exploratory cohort and the observed ones in the validation cohort. Overall, correlation analysis revealed highly significant relationships between the two datasets (p<0.001 for both treatments). In Bland-Altman analysis, mean bias was 0% in patients on amlodipine and 2% in those treated with valsartan without evidence of proportional bias.

**Figure 3.**
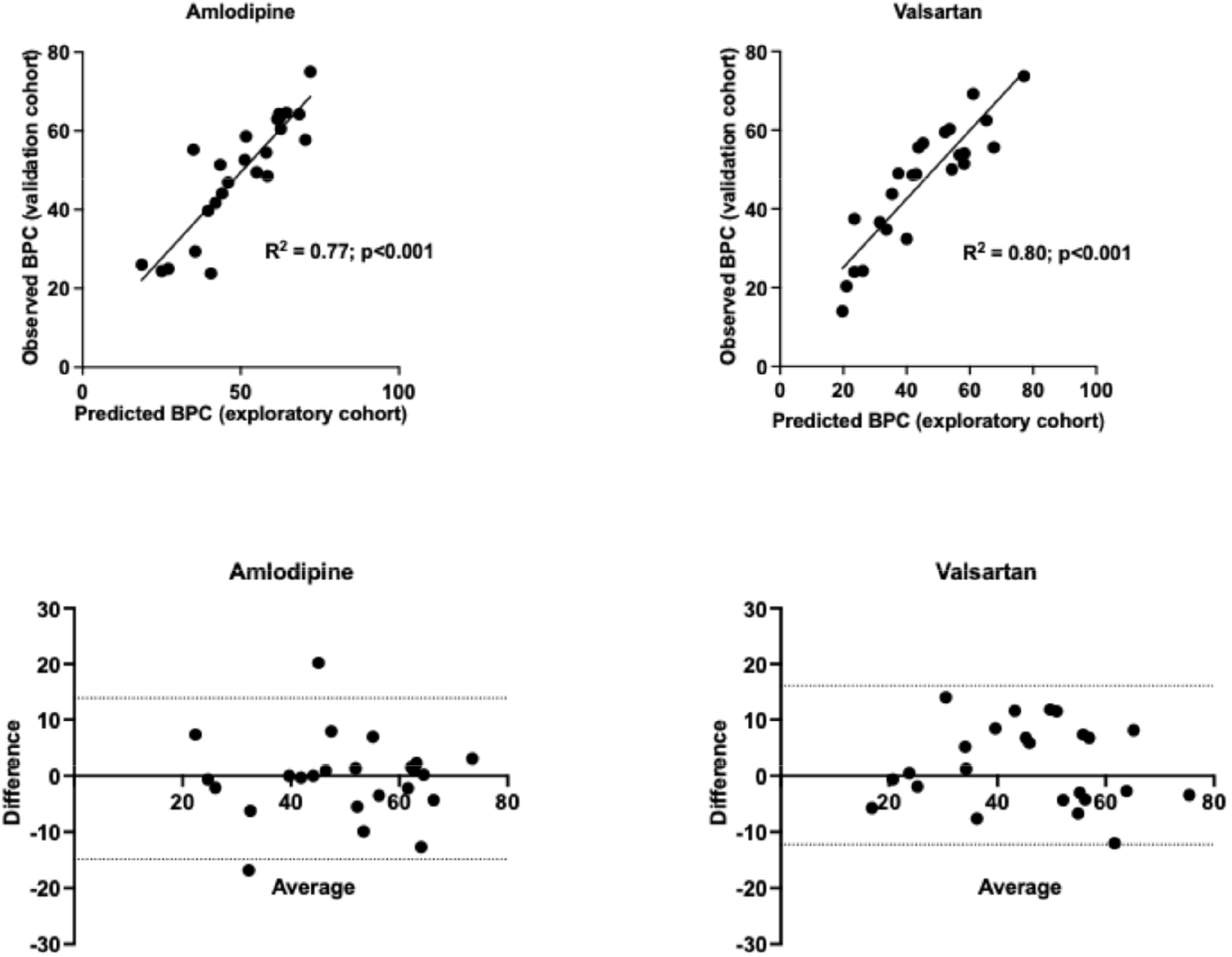
Upper panel: correlations between percentage of blood pressure control in the exploratory cohort (x-axis) and the observed percentage of blood pressure control in the validation cohort (y-axis). Lower panel: Bland-Altman plots of the same data.

Logistic regression analysis showed that in the validation cohort, 8 and 9 profiles from the amlodipine and the valsartan group respectively remained significantly associated with BP control. Although fewer profile-based associations reached statistical significance in the validation cohort, this is likely due to low numbers. In both treatment groups, a core set of 7 profiles (14, 62, 68, 86, 87, 92 and 95) showed reproducible effect sizes across the exploratory and the validation cohort.

When the two treatment groups were again combined, a similar pattern emerged as in the exploratory cohort.

### Profiles versus single predictors

Because diabetes has a profound influence on the effectiveness of antihypertensive medication,^11^ we explored in separate analyses whether profiling added any information over and above that conferred already by the presence or absence of diabetes. To this end, we compared a model with diabetes as single predictor to a model that included patient profiles. Likelihood ratio tests showed that both in diabetics and in non-diabetics, profiling predicted BP control better than diabetes status alone (p<0.001 for both). Remarkably, among diabetic patients, profiles 20, 23, 68 and 71 achieved significantly better-than-average BP control, whereas a worse response was seen in profiles 69, 72, 92 and 95. Among non-diabetics, on the other hand, significantly better-than-average responses were seen in profiles 14, 17, 50, 62 and 65 and worse responses in profiles 41, 63, 85, 86, 87, 89 and 134. It is noteworthy that five of the last seven profiles had no comorbid conditions at all.

As obesity may also reduce the therapeutic efficacy of antihypertensives,^12^ we performed a similar analysis using obesity (in non-diabetic patients) as the single stratifying factor. Again, profiling had a significant overall effect on BP control in both obese and non-obese patients (likelihood ratio test, p<0.001 for both groups). Profiling also predicted BP control better than age, baseline systolic BP and previous monotherapy as single factors. However, the treatment-naïve group and the group with two or more previous medications were too small to allow a meaningful analysis. Likewise, there were not enough patients without organ damage to test whether profiling performed better than organ damage alone.

### Association between patient profiles and cardiovascular outcome

In the exploration cohort, 1464 patients had a primary event against 712 in the validation cohort. Cox proportional hazard models demonstrated that profile categories were significantly associated with the primary endpoint of the trial (likelihood ratio test: p<0.001 in both groups). Although crude hazard ratios varied across the categories, by and large the direction and magnitude of the associations in the exploratory cohort were consistent between the two treatment groups. Women were less at risk than men at all profile categories (likelihood ratio test: p<0.001) but there was no interaction between sex and profile-related outcome.

In the validation cohort, the effect estimates aligned well with those in the exploratory cohort, but again less associations reached statistical significance. Therefore, for further analyses we combined the data from the two cohorts to increase power. Profile categories were independently associated with outcome. Eight profiles in the amlodipine group and ten in the valsartan group showed significant associations with the primary endpoint. All eight profiles in the amlodipine group were also associated with outcome in the valsartan group.

After Hommel’s correction, five and seven profiles remained in the amlodipine and valsartan group respectively (table 4). In both treatment groups, profile 14 which represents a relatively favourable condition, lowered the risk of an event, while profiles 69, 71and 95 increased the risk. Except for a similar age range, the latter three profiles share as common feature the presence of organ damage as well as diabetes. Although the direction of the associations remained when diabetics and non-diabetics were considered separately, most significances were lost due to a lack of power when both treatments were considered separately. After combining the two treatment groups, diabetics still did not exhibit any significant association between profiles and events, indicating that profiling does not add to the risk conferred by diabetes alone. In the non-diabetic group, on the other hand, five significant associations emerged. While profiles 14 (p<0.001) and 50 (p=0.02) correlated with a better-than-average outcome, profiles 66 (with obesity as comorbidity) and profiles 110 and 134 (both without comorbid conditions) were associated with a worse outcome (p-values 0.02, 0.02 and <0.001 respectively).

**Table 4.** Hazard ratios (with 95% confidence intervals) for cardiovascular events in relation to profile categories. Adjusted for sex.

| Treatment | Profile | HR | p (crude) | p (Hommel) | Treatment | Profile | HR | p (crude) | p (Hommel) |
| --- | --- | --- | --- | --- | --- | --- | --- | --- | --- |
| Amlodipine | 14 | 0.45 (0.27-0.76) | **0.003** | **0.01** | Valsartan | 14 | 0.28 (0.15-0.53) | **0.001** | **0.001** |
| Amlodipine | 17 | 0.75 (0.44-1.28) | 0.29 | 0.47 | Valsartan | 17 | 0.79 (0.46-1.35) | 0.38 | 0.51 |
| Amlodipine | 20 | 1.47 (0.84-2.57) | 0.18 | 0.33 | Valsartan | 20 | 1.02 (0.56-1.88) | 0.94 | 0.94 |
| Amlodipine | 23 | 1.02 (0.59-1.79) | 0.93 | 0.97 | Valsartan | 23 | 1.45 (0.88-2.39) | 0.15 | 0.27 |
| Amlodipine | 38 | 0.59 (0.29-1.22) | 0.15 | 0.32 | Valsartan | 38 | 0.91 (0.50-1.67) | 0.76 | 0.80 |
| Amlodipine | 41 | 0.81 (0.37-1.77) | 0.60 | 0.76 | Valsartan | 41 | 0.61 (0.20-1.81) | 0.37 | 0.51 |
| Amlodipine | 50 | 0.13 (0.02-0.86) | **0.03** | 0.11 | Valsartan | 50 | 0.20 (0.05-0.77) | **0.02** | 0.06 |
| Amlodipine | 56 | 0.92 (0.44-1.88) | 0.81 | 0.88 | Valsartan | 56 | 0.75 (0.32-1.75) | 0.51 | 0.64 |
| Amlodipine | 62 | 0.82 (0.64-1.05) | 0.11 | 0.27 | Valsartan | 62 | 0.76 (0.60-0.97) | **0.02** | 0.06 |
| Amlodipine | 63 | 0.81 (0.51-1.31) | 0.39 | 0.56 | Valsartan | 63 | 0.93 (0.61-1.41) | 0.72 | 0.80 |
| Amlodipine | 65 | 1.01 (0.72-1.41) | 0.97 | 0.97 | Valsartan | 65 | 0.82 (0.57-1.17) | 0.27 | 0.44 |
| Amlodipine | 66 | 1.13 (0.61-2.07) | 0.70 | 0.80 | Valsartan | 66 | 1.61 (0.99-2.61) | 0.05 | 0.11 |
| Amlodipine | 68 | 1.40 (1.02-1.86) | **0.04** | 0.11 | Valsartan | 68 | 1.44 (1.08-1.94) | **0.01** | **0.05** |
| Amlodipine | 69 | 2.12 (1.34-3.37) | **0.001** | **0.01** | Valsartan | 69 | 1.89 (1.18-3.02) | **0.008** | **0.03** |
| Amlodipine | 71 | 1.85 (1.36-2.51) | **<0.001** | **0.001** | Valsartan | 71 | 1.90 (1.42-2.55) | **<0.001** | **<0.001** |
| Amlodipine | 72 | 1.86 (1.11-3.13) | **0.02** | 0.08 | Valsartan | 72 | 2.04 (1.29-3.22) | **0.002** | **0.01** |
| Amlodipine | 85 | 0.68 (0.36-1.29) | 0.24 | 0.40 | Valsartan | 85 | 0.70 (0.38-1.29) | 0.25 | 0.43 |
| Amlodipine | 86 | 1.07 (0.81-1.39) | 0.64 | 0.77 | Valsartan | 86 | 0.74 (0.54-1.01) | 0.06 | 0.11 |
| Amlodipine | 87 | 1.22 (0.74-2.02) | 0.43 | 0.58 | Valsartan | 87 | 0.92 (0.54-1.58) | 0.77 | 0.80 |
| Amlodipine | 89 | 0.54 (0.29-1.03) | 0.06 | 0.26 | Valsartan | 89 | 1.08 (0.71-1.64) | 0.73 | 0.80 |
| Amlodipine | 92 | 1.18 (0.83-2.98) | 0.36 | 0.54 | Valsartan | 92 | 1.57 (1.16-2.12) | **0.003** | **0.02** |
| Amlodipine | 95 | 2.12 (1.56-2.98) | **<0.001** | **0.001** | Valsartan | 95 | 1.83 (1.26-2.65) | **0.002** | **0.01** |
| Amlodipine | 110 | 1.56 (0.44-2.85) | 0.16 | 0.32 | Valsartan | 110 | 1.34 (0.75-2.39) | 0.32 | 0.48 |
| Amlodipine | 134 | 2.35 (1.34-4.10) | **0.003** | **0.01** | Valsartan | 134 | 1.88 (1.01-3.29) | **0.03** | 0.06 |

HR: hazard ratio. Significant effects indicated in bold. P (crude): p-value in crude analysis. P (Hommel): p-value after Hommel's correction for multiple testing.

Figure 4 shows the relationship between the percentage BP control and the corresponding absolute event rate for each profile. When both treatment groups were combined, a significant inverse relationship was apparent. However, there is substantial variability in the data and only 14% of the variation in event rate is explained by BP control.

**Figure 4.**
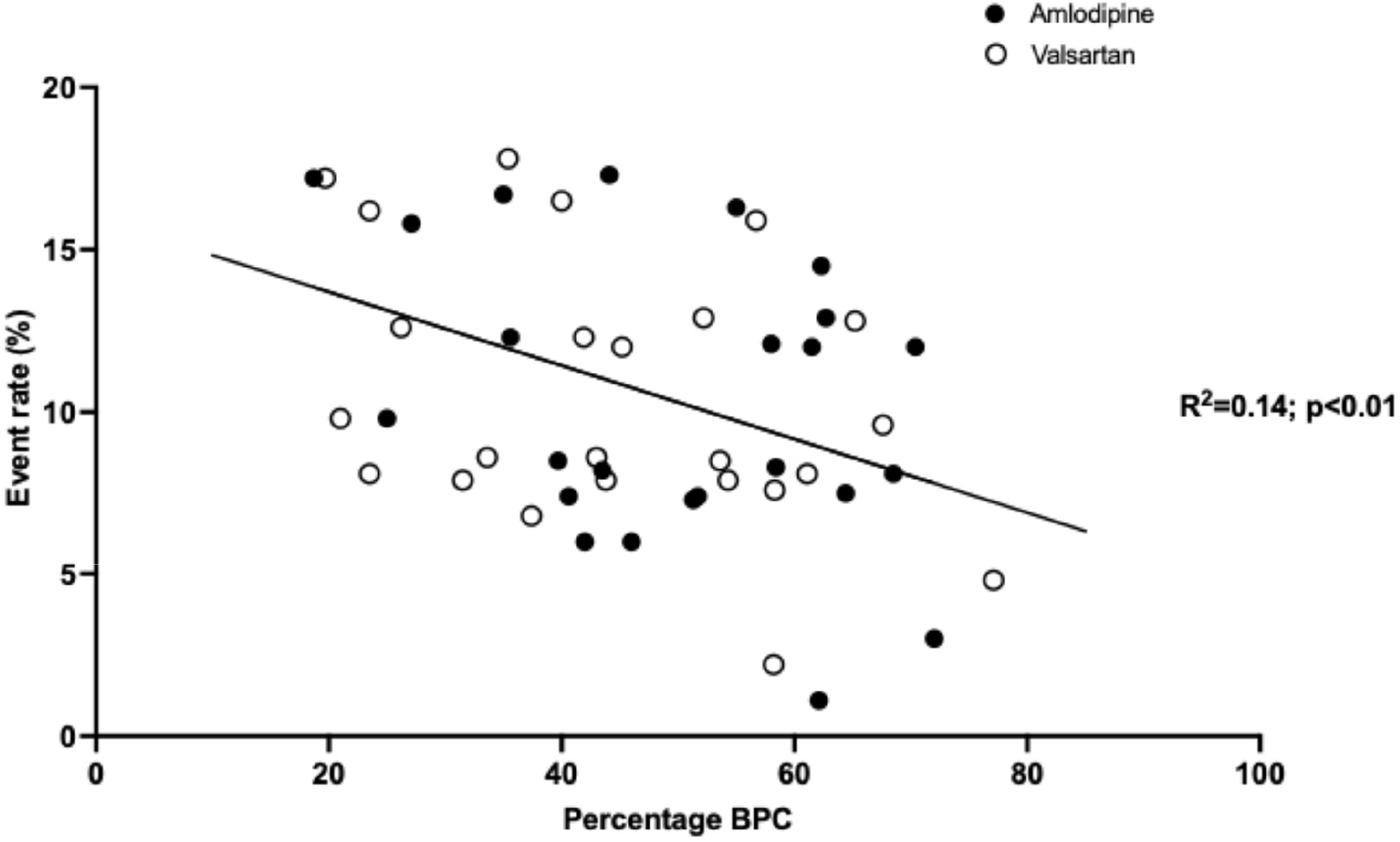
Relationship between percentage blood pressure control and event rates. Each data point represents one profile. BPC: blood pressure control. Closed circles: amlodipine; open circles: valsartan.

### Adverse events

The proportion of patients with an adverse event of any kind was low and, with the exception of peripheral oedema, favored amlodipine. However, no associations whatsoever were found between the various profiles and the incidence of adverse events, irrespective of whether the two treatment groups were considered separately or in combination.

## Discussion

In the present exploratory analysis of the VALUE trial, we identified several patient profiles that were significantly associated with the binary outcome BP control at six months or at the end of follow-up. The majority of these associations in the exploratory cohort were also found in the validation cohort with consistent effect sizes among both treatment groups. The small differences that we observed in the validation cohort are likely due to smaller numbers rather than to pathophysiological or pharmacodynamic factors.

The identification of a core set of profiles which can consistently be linked to a reduced BP response as well as the absence of a significant interaction between profile categories and treatment group suggest that the effect of treatment on BP is mainly determined by specific patient profiles, even in patients with diabetes. From a clinical perspective, this is important because it implies that for hypertensive patients a profile-based therapeutic regimen may be more important than treatment based on a particular drug type. Indeed, several profiles that were found to be unfavorable in the present study were also associated with worse BP control in the INSIGHT study in which other medications were used.^3^ Consequently, our approach of patient profiling may also be applicable to other antihypertensive strategies.

The fact that profiles were not significantly associated with adverse events adds to the hypothesis that patient profiling correlates more closely with prognostic factors rather than with the specific pharmacological effects of treatment. Although our data align well with the recommendations of the Prognosis Research Strategy (PROGRESS) group,^13^ prospective trials are needed to confirm this hypothesis. If confirmed, this may call for more aggressive therapeutic strategies in patients with high-risk profiles such as those found in the present study.

Except for BP control, profiles were also related to the primary endpoint of the trial. However, diabetes is in itself such an overriding risk factor that profiling does not have any additional value for estimating cardiovascular risk in patients with this disease. In non-diabetics, on the other hand, we identified three profiles with an adverse outcome. Remarkably, two of these profiles did not contain comorbidity. This suggests that profiling could be particularly useful in non-diabetics to select those patients in whom more aggressive treatment may be warranted.

Given the robust association between profile categories and BP control, it is tempting to speculate that the effect of the various profiles on outcome is mediated by the degree of BP control in those profiles. The inverse relationship between the percentage BP control and event rates certainly lends support to that hypothesis. However, this relationship is not particularly strong and indicates that BP control alone cannot explain the association between profiles and events. This suggests that profile classification captures elements of cardiovascular risk that go beyond BP control per se. This was further corroborated when we examined the effect of sex. We found that for the same profiles and, hence, a comparable responsiveness to treatment, women were less at risk than men. Any sex-related difference in cardiovascular risk must, therefore, be due to factors other than BP control. Altogether, our data fuel the idea of specific pathways being linked to certain patient profiles that carry prognostic, BP independent, information. Whether this involves, for instance, arterial stiffness, endothelial dysfunction or other pathways cannot be deducted from the present data. However, recognition of such pathways may help us to further improve the management of hypertension and to reduce so-called residual risk.^14^

Our results should be interpreted within the context of several limitations. First, our study is an exploratory analysis of existing data and not based on a randomized treatment design. Therefore, our data should be considered as hypothesis-generating and not as firm evidence. Second, while internal validation was successful, external validation in independent populations remains mandatory to confirm generalizability, although the similarity with the

INSIGHT data is reassuring in this respect. Third, the number of patients per profile differed considerably with some profiles being represented by relatively small sample sizes. This may have led to unstable estimates and reduced power in the validation cohort. It would be worthwhile, therefore, to repeat the analyses in a much larger group of patients. Finally, we have to acknowledge that our analysis was based on only 24 of the 144 patient profiles that were theoretically possible with our approach. Therefore, the inevitable conclusion is that for the majority of profiles no compelling information is available yet regarding the effects of treatment. Future research should focus on external validation of our results in a large group of other trials and on the potential role of profiling in clinical decision making when it comes to the treatment of hypertensive patients. Also, more extensive profiling with the addition of other prognostic markers needs to be explored.

## Data Availability

Data will be made available upon reasonable request following publication.

## References

1. Sundström J, Lind L, Nowrouzi S, Hagstrom E, Held C, Lytsy P, Neal B, Marttala K, Ostlund O. Heterogeneity in Blood Pressure Response to 4 Antihypertensive Drugs: A Randomized Clinical Trial. JAMA. 2023;329:1160–1169. doi: 10.1001/jama.2023.3322

2. Shang Q, Yao S, Ouyang M, Wang X, Luo S. Secondary Analysis of Randomized Controlled Trials: Methodological Considerations and Best Practices. Ann Clin Epidemiol. 2025;7:137–147. doi: 10.37737/ace.25017

3. De Leeuw PW, Mancia G, Palmer CR, Ruilope LM, Schalm SW, Brown MJ. Patient-profiled treatment responses in a large hypertension trial: a posthoc analysis of the INSIGHT study. J Hypertens. 2026;444:999–1004. doi: 10.1097/HJH.0000000000004284

4. Julius S, Kjeldsen SE, Weber M, Brunner HR, Ekman S, Hansson L, Hua T, Laragh J, McInnes GT, Mitchell L, et al. Outcomes in hypertensive patients at high cardiovascular risk treated with regimens based on valsartan or amlodipine: the VALUE randomised trial. Lancet. 2004;363:2022–2031.

5. Weber MA, Julius S, Kjeldsen SE, Brunner HR, Ekman S, Hansson L, Hua T, Laragh JH, McInnes GT, Mitchell L, et al. Blood pressure dependent and independent effects of antihypertensive treatment on clinical events in the VALUE Trial. Lancet. 2004;363:2049–2051.

6. Cushman WC, Ford CE, Einhorn PT, Wright JT, Jr., Preston RA, Davis BR, Basile JN, Whelton PK, Weiss RJ, Bastien A, et al. Blood pressure control by drug group in the Antihypertensive and Lipid-Lowering Treatment to Prevent Heart Attack Trial (ALLHAT). J Clin Hypertens (Greenwich). 2008;10:751–760. doi: 10.1111/j.1751-7176.2008.00015.x

7. Mann J, Julius S, for the VALUE Trial Group. The Valsartan Antihypertensive Long-term Use Evaluation (VALUE) trial of cardiovascular events in hypertension. Rationale and design. Blood Press. 1998;7:176–183.

8. Oldham PD. A note on the analysis of repeated measurements of the same subjects. J Chronic Dis. 1962;15:969–977.

9. Gill JS, Zezulka AV, Beevers DG, Davies P. Relation between initial blood pressure and its fall with treatment. Lancet. 1985;1:567–569. doi: 10.1016/s0140-6736(85)91219-x

10. Hommel G. A stagewise rejective multiple test procedure based on a modified Bonferroni test. Biometrika. 1988;75 (2):383–386.

11. Gnanenthiran SR, Webster R, Silva A, Maulik PK, Salam A, Selak V, Guggilla RK, Schutte AE, Patel A, Rodgers A, et al. Reduced efficacy of blood pressure lowering drugs in the presence of diabetes mellitus-results from the TRIUMPH randomised controlled trial. Hypertens Res. 2023;46:128–135. doi: 10.1038/s41440-022-01051-7

12. Bhandari P, Prakash V, Flack JM. Influence of Obesity on Blood Pressure Responses to Antihypertensive Drug Therapy in an Urban Hypertension Specialty Clinic. Am J Hypertens. 2022;35:740–744. doi: 10.1093/ajh/hpac072

13. Riley RD, Hayden JA, Steyerberg EW, Moons KG, Abrams K, Kyzas PA, Malats N, Briggs A, Schroter S, Altman DG, et al. Prognosis Research Strategy (PROGRESS) 2: prognostic factor research. PLoS Med. 2013;10:e1001380. doi: 10.1371/journal.pmed.1001380

14. Manta E, Thomopoulos C, Kariori M, Polyzos D, Mihas C, Konstantinidis D, Farmakis D, Mancia G, Tsioufis K. Revisiting Cardiovascular Benefits of Blood Pressure Reduction in Primary and Secondary Prevention: Focus on Targets and Residual Risk-A Systematic Review and Meta-Analysis. Hypertension. 2024;81:1076–1086. doi: 10.1161/HYPERTENSIONAHA.123.22610

